# p-tau/Aβ42 Ratio Associates with Cognitive Decline in Alzheimer’s disease, Mild Cognitive Impairment, and Cognitively Unimpaired Older Adults

**DOI:** 10.1101/2020.10.13.20211375

**Authors:** Ruchika Shaurya Prakash, Michael R. McKenna, Oyetunde Gbadeyan, Rebecca Andridge, Douglas W. Scharre, for the Alzheimer’s Disease Neuroimaging Initiative

**Author notes:** Data used in preparation of this article were obtained from the Alzheimer’s Disease Neuroimaging Initiative (ADNI) database (adni.loni.usc.edu). As such, the investigators within the ADNI contributed to the design and implementation of ADNI and/or provided data but did not participate in analysis or writing of this report. A complete listing of ADNI investigators can be found at: http://adni.loni.usc.edu/wp-content/uploads/how_to_apply/ADNI_Acknowledgement_List.pdf. Correspondence concerning this article should be addressed to Ruchika Shaurya Prakash, Department of Psychology, The Ohio State University, 139 Psychology Building, 1835 Neil Avenue, Columbus, OH 43210. **Disclosures** R Prakash reports no disclosures. M McKenna reports no disclosures. O. Gbadeyan reports no disclosures. R. Andridge reports no disclosures. D. Scharre reports no disclosures.

## Abstract

**INTRODUCTION:** The most well-studied biomarkers in AD are CSF amyloid beta-42 (Aβ_42_), tau, p-tau, and the ratio p-tau/Aβ_42_. The ratiometric measure of p-tau/Aβ_42_ shows the best diagnostic accuracy, and correlates reliably with metrics of cognition in unimpaired participants. However, no study has examined the impact of the CSF p-tau/Aβ_42_ ratio in predicting cognitive decline in both healthy and AD individuals in one sample. The goal of this study was to examine whether CSF-based p-tau/Aβ_42_ predicts changes in global cognitive functioning, episodic memory, and executive functioning over a two-year period in cognitively impaired older adults (CU), and in individuals with Mild Cognitive Impairment (MCI) and Alzheimer’s disease (AD).

**METHODS:** This study involves secondary analysis of data from 1215 older adults available in the Alzheimer’s Disease Neuroimaging Initiative (ADNI). Neuropsychological variables, collected at baseline, 6-month, 12-month, and 24-month follow-ups, included the Preclinical Alzheimer’s Cognitive Composite (PACC) to assess global cognitive functioning, ADNI-MEM to assess episodic memory functioning, and ADNI-EF to assess executive functioning. Linear mixed models were constructed to examine the effect of CSF p-tau/Aβ_42_, diagnostic group, and change over time (baseline, 6-month, 12-month, and 24-month) on cognitive scores.

**RESULTS:** CSF p-tau/Aβ_42_ ratios predicted worsening cognitive impairment, both on global cognition and episodic memory in individuals with MCI and AD, but not in CU older adults and predicted decline in executive functioning for all three diagnostic groups.

**DISCUSSION:** Our study, including CU, MCI, and AD individuals, provides evidence for differential cognitive consequences of accumulated AD pathology based on diagnostic groups.

## Introduction

There has been an increased global focus in understanding the etiology and treatment possibilities for Alzheimer’s disease (AD). AD has been traditionally conceptualized as a clinical pathologic syndrome, with multi-domain cognitive symptoms, considered the defining feature of the disease^1–3^. However, accumulating evidence from imaging and autopsy studies evince support for the onset of pathophysiological processes well before the onset of AD symptoms^4,5^, and in some instances, without the presence of known cognitive symptoms^6–9^, thus reinforcing a revised conceptualization of the disease away from a syndromal manifestation to a biological definition^10,11^. This redefinition of AD as a neuropathological disease prioritizes the investigation of misfolded and aggregated amyloid beta (Aβ) peptides and hyperphosphorylated tau proteins, detected in-vivo using either CSF- or PET-based examination, for the understanding of AD and its clinical manifestation.

Cerebral Aβ accumulation can be estimated through a reduction in Aβ_42_ concentration, one of the most reliable diagnostic isoforms of Aβ, in CSF^12^ derived from a lumbar puncture. Importantly, reduction in CSF Aβ_42_ has been observed in cognitively unimpaired (CU) older adults, with clinicopathological studies evincing support for amyloid pathology as one of the earliest detectable markers of AD pathology in living persons^13–15^. However, β-amyloidosis does not perfectly predict the clinical expression of AD, with tauopathy essential for the manifestation of cognitive deterioration observed in AD^11,16,17^. Two of the most thoroughly examined CSF-based tau biomarkers are phosphorylated tau (p-tau) and total tau (t-tau) known to be made up of various protein isomers^18^. P-tau concentration is thought to be more specific to AD pathology, reflecting the hyperphosphorylated tau found in neurofibrillary tangles in the brain and correlating with the severity of paired helical filament tau aggregation at autopsy ^19,20^, whereas t-tau concentration is thought to reflect general neurodegeneration and can be elevated in other neurological disorders like traumatic brain injury and stroke^21,22^.

Of these CSF-based markers, the ratio CSF tau/Aβ_42_, combining variance across the two critically implicated proteinopathies of Aβ and tau, provides one of the best diagnostic accuracies among fluid- and PET imaging-based biomarkers^23–26^. For example, Hansson et al. compared the accuracy rates for Elecsys-based immunoassays of Aβ_42_, t-tau/Aβ_42_, and p-tau/Aβ_42_, and showed that the two ratio measures have stronger concordance with amyloid PET than CSF Aβ_42_ alone^27^. This ratio allows us to capture a biologic measure of the two most prominent neuropathological features of AD creating high specificity for diagnostic classification of the disease.

Despite there being a shift from the traditional clinical-pathological conceptualization of AD emphasizing clinical consequences of the disease to a more biological definition highlighting underlying pathological processes, the two are inextricably linked, with studies systematically investigating associations between molecular and clinical changes. Although there have been studies reporting significant associations between accumulation of amyloid pathology and metrics of global cognition, episodic memory, and executive functioning^28,29^, there have also been studies reporting weaker correlations between CSF-Aβ_42_ and cognitive functioning^30,31^. A handful of studies, notably, have examined the combined contribution of amyloidosis and tauopathy, and evince support for these models to best predict variance in baseline cognitive functioning and trajectory of decline in cognitive functioning^32,33^. Given the reconceptualization of AD as a biological disease, with accumulation of misfolded proteins occurring decades before the onset of cognitive sequalae, this study examined the interaction between diagnostic status, as defined in the Alzheimer’s Disease Neuroimaging Initiative (ADNI) and the ratio of CSF p-tau/Aβ_42_, in predicting changes in cognitive functioning over a two-year period.

## Methods

Data were obtained from the ADNIMERGE R package on March 27, 2020. ADNI is an ongoing, multi-site, longitudinal study designed to examine the role of fluid-based and imaging-based biomarkers in healthy and pathological aging. Ongoing since 2003, the study has had four waves of data collection (ADNI-1, ADNI-GO, ADNI-2, and ADNI-3), with baseline data from 2250 participants publicly available. In this study, anyone with available baseline CSF biomarker data from the ADNI1, ADNI-GO and ADNI-2 were included in the analysis, resulting in a total of 1215 participants.

### Participants

We included ADNI CU older adults, ADNI MCI, and ADNI AD subjects in this study. General inclusion criteria for the ADNI study included: ages between 55-90 years, English or Spanish speaking, Hachinski Ischemic Score of less than or equal to 4, adequate visual and auditory acuity, good general health, at least 6 years of education or equivalent work history, and a Geriatric Depression Scale score of less than 6.

Diagnostic criteria for classifying participants at baseline involved a combination of subjective reports, neuropsychological assessment, and physician assessment^34^. Specifically, CU participants had to meet the following criteria: 1) Mini-Mental Status Examination (MMSE) score ≥24, 2) scoring above education-adjusted cut-offs on the delayed free recall of the Logical Memory II subscale (≥9 for 16+ years of education, ≥5 for 8-15 years of education, ≥3 for 0-7 years of education), 3) a Clinical Dementia Rating score of 0, and 4) absence of AD dementia or any other neurological condition. Individuals with subjective memory concerns (SMC) had the same diagnostic criteria as CU individuals, except reported a significant subjective memory concern as indexed by the Cognitive Change Index; a self-report measure where participants are asked to compared present cognitive functioning with the last five years. The first 12 questions focus on memory concerns, and older adults who scored ≥ 16 on the first 12 questions were classified as SMC. As the only difference between CU and SMC was self-reported assessment of declines in memory functioning, we combined these two groups for this study. For MCI participants, the following criteria were employed: 1) MMSE score ≥ 24, 2) scoring within education-adjusted range for the Logical Memory II subscale (EMCI: 9-11 for 16+ years of education, 5-9 for 8-15 years of education, 3-6 for 0-7 years of education; LMCI: ≤8 for 16+ of education, ≤4 for 8-15 years of education, ≤2 for 0-7 years of education), 3) a Clinical Dementia Rating score of 0.5, 4) self or partner reported memory complaint(s), and 5) absence of AD dementia or any other neurological condition. Given that the only difference between EMCI and LMCI participants was performance on the Logical Memory II subscale, these two groups were combined for the purpose of the current study. AD participants were required to meet the following criteria: 1) MMSE score between 20-26, 2) scoring below education-adjusted cut-offs for the Logical Memory Scale II (8 for 16+ of education, ≤4 for 8-15 years of education, ≤2 for 0-7 years of education), 3) a Clinical Dementia Rating score of 0.5 or 1.0, and 4) self or partner reported memory complaints.

Of the 1215 participants included in the study, 367 were classified as CU, 619 were classified as MCI, and 229 were classified as AD. Table 1 presents the demographic and relevant clinical characteristics for each of the three groups. Due to the low presence of double ε4 alleles in the CU sample, participants with single and double ε4 alleles were collapsed into one group “*APOE* ε4 present.”

**Table 1:** Baseline characteristics of participants by diagnostic group

| Characteristic | CU (n=367) |  | MCI (n=619) |  | AD (n=229) |  | P-value |
| --- | --- | --- | --- | --- | --- | --- | --- |
|  | Mean (SD) or N (%) | Range | Mean (SD) or N (%) | Range | Mean (SD) or N (%) | Range |  |
| Sex |  |  |  |  |  |  | 0.001 |
| Female | 193 (53%) |  | 255 (41%) |  | 95 (41%) |  |  |
| Male | 174 (47%) |  | 364 (59%) |  | 134 (59%) |  |  |
| Race* |  |  |  |  |  |  | 0.03 |
| White | 333 (91%) |  | 583 (94%) |  | 219 (96%) |  |  |
| American Indian / Alaskan | 1 (0.3%) |  | 1 (0.2%) |  | 0 (0%) |  |  |
| Asian | 4 (1%) |  | 9 (1%) |  | 4 (2%) |  |  |
| Black | 24 (7%) |  | 15 (2%) |  | 5 (2%) |  |  |
| Hawaiian / Pacific Islander | 0 (0%) |  | 2 (0.3%) |  | 0 (0%) |  |  |
| More than One Race | 5 (1%) |  | 7 (1%) |  | 1 (0.5%) |  |  |
| Age (years) | 73.8 (5.9) | 56.2 to 89.6 | 72.3 (7.5) | 54.4 to 91.4 | 74.6 (8.2) | 55.6 to 90.3 | <.0001 |
| Years of education | 16.4 (2.6) | 6 to 20 | 16.1 (2.8) | 6 to 20 | 15.4 (2.9) | 4 to 20 | 0.0004 |
| *APOE* |  |  |  |  |  |  | <.0001 |
| ε4 allele absent | 264 (72%) |  | 314 (51%) |  | 73 (32%) |  |  |
| 1 ε4 allele | 94 (26%) |  | 239 (39%) |  | 107 (47%) |  |  |
| 2 ε4 alleles | 9 (2%) |  | 66 (11%) |  | 49 (21%) |  |  |
| *APOE*, collapsed |  |  |  |  |  |  | <.0001 |
| ε4 allele absent | 264 (72%) |  | 314 (51%) |  | 73 (32%) |  |  |
| ε4 allele(s) present | 103 (28%) |  | 305 (49%) |  | 156 (68%) |  |  |

CU = Cognitively unimpaired; MCI = Mild cognitive impairment; AD = Alzheimer's disease
*APOE* = apolipoprotein E genotype
\*Race missing for 2 subjects in the MCI group

### Neuropsychological Measures and Composites

Validated neuropsychological composites of global cognition, episodic memory, and executive functioning were employed to examine the impact of AD pathology on two-year (baseline, 6-month, 12-month, and 24-month) changes in key cognitive domains impacted in AD. For global cognition, we employed the Preclinical Alzheimer’s Cognitive Composite (PACC)^35^, assessing domains of episodic memory, executive functioning, and overall cognition. PACC includes the following measures: MMSE total score, Trails-Making Test B, delayed recall score from the Logical Memory II subscale, and the delayed word recall from the Alzheimer’s Disease Assessment Scale–Cognitive Subscale (ADAS-COG).

ADNI-MEM was employed to index changes in memory, and includes performance on the Logical Memory I and II tasks, several item scores on the Rey Auditory Verbal Learning Test, the cognitive subscale of the Alzheimer’s Disease Assessment Scale, and the three word recall items from the MMSE^36^. Similarly, to measure executive functioning, we employed the ADNI-EF made up of scores on the Digit Symbol Substitution test from the Weschler Adult Intelligence Scale-Revised, Digit Span Backwards Test, Trails-Making A and B, Category Fluency, and Clock Drawing^37^.

Table 1 presents the data available at each time point for the three diagnostic groups. Although ADNI has longer-term follow-up data, there is significant attrition as a function of the diagnostic group, such that at 36 months, only 23 participants with AD, 441 with MC, and 132 with CU and baseline CSF data had the composite measure of EF. As such, we restricted our analysis to a two-year follow-up.

### CSF Biomarker Assessment and Classification

CSF biomarker concentrations were pulled from the upennbiomk9.rdata file nested in the ADNIMERGE R package. P-tau and Aβ_42_ concentrations were measured in picograms per milliliter (pg/mL) by ADNI researchers using the highly automated Roche Elecsys immunoassays on the Cobas e601 automated system. Of the 1215 participants, 191 participants met the upper limit for the Aβ_42_ concentration (1700 pg/ml for the Roche-based assaying), and we employed the extrapolated values provided by the ADNI group. For all participants, we computed the ratio of p-tau/Aβ_42_, with higher values representing higher pathological state of the two proteinopathies. Additionally, to quantify the distribution of clinically significant AD pathology in the three diagnostic groups, we employed the cut-off of .028 provided by Hansson et al. ^27^ for the ADNI dataset showing concordance of 91.8% with standardized uptake value ratio for amyloid-β PET and 90.3% with visual read within the ADNI sample.

### Statistical Analysis

Comparisons across diagnostic groups on baseline measures were made using ANOVA and chi-square tests as appropriate. Changes in outcomes (PACC, ADNI-MEM, ADNI-EF) over time were modeled using linear mixed effects models. An advantage of this method is that subjects who may be missing measurements at some time points can still contribute to the analysis. Initial exploratory analysis revealed that the trajectories of all three outcomes were linear, and thus time was included in the models as a linear effect. In the first set of models, fixed effects included time (months; linear), diagnostic group (CU, AD, MCI), and their interaction. Additional covariates included to guard against confounding were age (at baseline), years of education, sex, and *APOE* status (ε4 allele present vs. absent). In the second set of models, baseline p-tau/Aβ_42_ ratio was added, including all two- and three-way interactions of tau/Aβ_42_ with time and diagnosis group, in order to test whether changes in cognitive measures over time were moderated by pathology. To capture the within-subject correlation arising due to repeated measurements over time, an unstructured residual error covariance was used for random subject-level error. The Kenward-Roger adjustment to the degrees of freedom was used to control Type 1 error.

Pairwise comparisons between groups were made using contrasts within these models.

### Data Availability

All data used in the current study have been downloaded from the publicly available Alzheimer’s Disease Neuroimaging Initiative database. We will also provide the curated data analyzed in the current study upon request by qualified investigators.

## Results

### Demographics Across Diagnostic Groups

As shown in **Table 1**, there were statistically significant differences for all demographic characteristics across diagnostic groups (*p* < .05 for all characteristics). Participants in the MCI and AD were more likely to be male and slightly more likely to be White. There were also small, but statistically significant differences in mean age and years of education across groups, with the MCI group having the youngest average age and the AD group having the lowest average number of years of education. There was also a large difference in *APOE* status across groups, with the ε4 allele(s) present in 68% of the AD group, 50% of the MCI group, and 28% of the CU group.

### Distribution of Pathology Across Diagnostic Groups

Aβ_42_, p-tau, and the p-tau/Aβ_42_ ratio differed significantly across the diagnostic groups (**Table 2**). Going from CU to MCI to AD, mean Aβ_42_ decreased, p-tau increased, and the p-tau/Aβ_42_ ratio increased. p-tau/Aβ_42_ ratio explained a larger amount of between-group variability (*R*^*2*^ = 0.20) compared to Aβ_42_ (*R*^*2*^ = 0.13) or p-tau (*R*^*2*^ = 0.12). **Figure 1** shows the distribution of the three AD biomarkers by group.

**Table 2:** Summary of pathology markers and cognitive measures by diagnostic group

| Measure | Diagnostic Group | Mean (SD) or N (%) | Range | P-value | R<sup>2</sup> |
| --- | --- | --- | --- | --- | --- |
| Aβ<sub>42</sub> | CU | 1336 (650) | 200 to 3592 | <.0001 | 0.13 |
|  | MCI | 1018 (555) | 210.9 to 3331 |  |  |
|  | AD | 693 (417) | 212.3 to 3139 |  |  |
| p-tau | CU | 21.8 (9.3) | 8 to 60 | <.0001 | 0.12 |
|  | MCI | 27.8 (15.0) | 8.2 to 120 |  |  |
|  | AD | 36.6 (15.8) | 8 to 95 |  |  |
| p-tau/Aβ<sub>42</sub> | CU | 0.022 (0.019) | 0.007 to 0.18 | <.0001 | 0.20 |
|  | MCI | 0.038 (0.031) | 0.006 to 0.24 |  |  |
|  | AD | 0.063 (0.034) | 0.008 to 0.25 |  |  |
| p-tau/Aβ<sub>42</sub> > 0.028 | CU | 86 (23%) | | <.0001 | -- |
|  | MCI | 319 (52%) |  |  |  |
|  | AD | 203 (89%) |  |  |  |
| Baseline PACC | CU | -0.26 (2.6) | -9.0 to 5.3 | <.0001 | 0.67 |
|  | MCI | -5.6 (3.9) | -16.0 to 2.7 |  |  |
|  | AD | -14.4 (3.1) | -23.0 to -4.7 |  |  |
| Baseline ADNI-MEM | CU | 1.05 (0.57) | -0.37 to 3.1 | <.0001 | 0.52 |
|  | MCI | 0.20 (0.69) | -1.5 to 2.3 |  |  |
|  | AD | -0.87 (0.52) | -2.8 to 0.57 |  |  |
| Baseline ADNI-EF | CU | 0.80 (0.82) | -1.3 to 3.0 | <.0001 | 0.31 |
|  | MCI | 0.23 (0.89) | -2.3 to 3.0 |  |  |
|  | AD | -0.90 (0.92) | -3.0 to 2.4 |  |  |

CU = Cognitively unimpaired; MCI = Mild cognitive impairment; AD = Alzheimer's diseases

**Figure 1:**
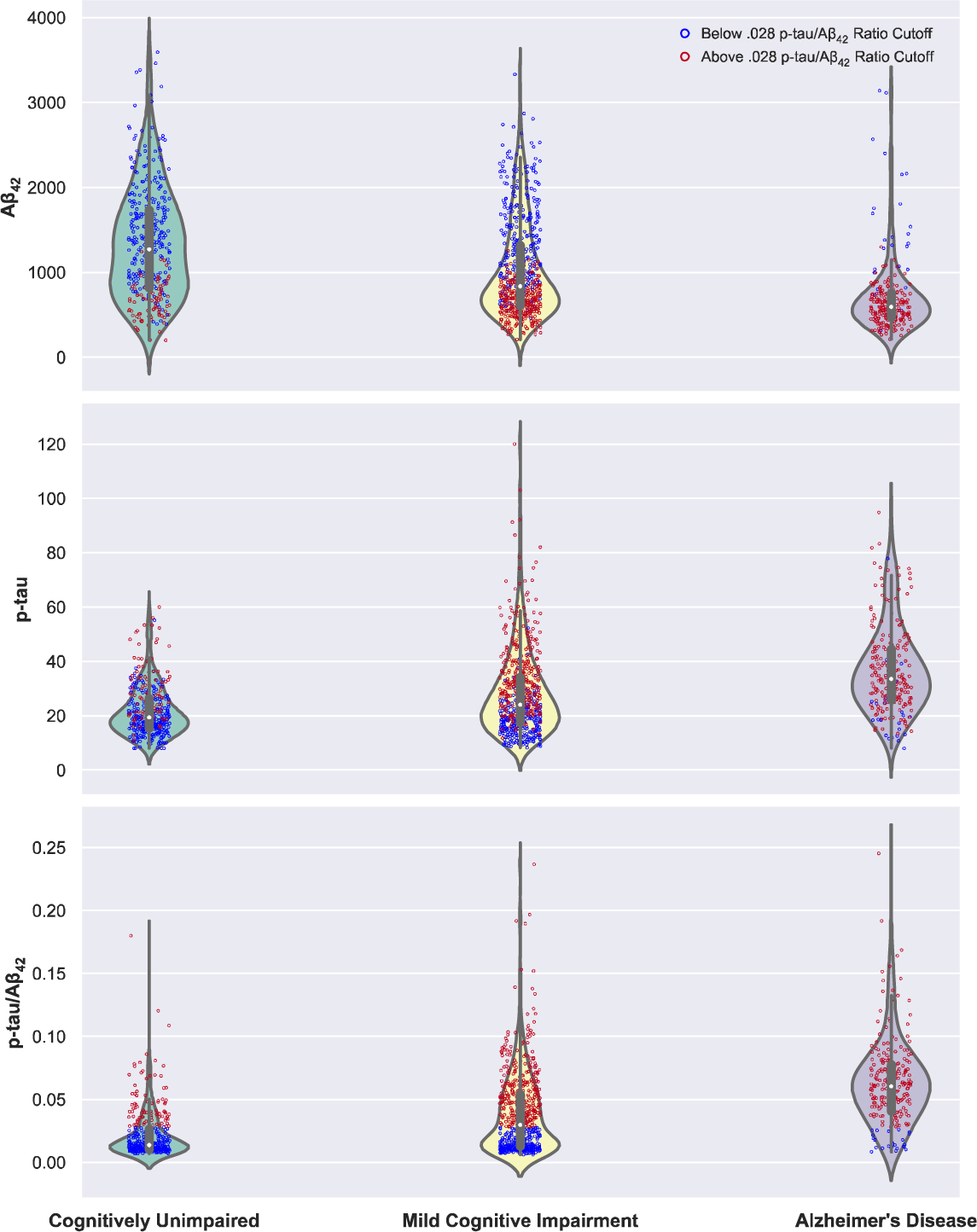
Presents the distribution of Aβ_42,_ p-tau, and the ratio of p-tau/Aβ_42_ across the three diagnostic groups. The individual data points are color coded with red dots for participants having a p-tau/Aβ_42_ value above the cut-off, and blue dots representing participants below the cut-off.

### Change in Cognitive Functioning Across Diagnostic Groups

At baseline there were large differences between groups on all three cognitive composites, with the AD group having lowest mean PACC, ADNI-MEM, and ADNI-EF and the CU group having the highest means (**Table 2; Figure 2a**). There were significant differences in the rate of change over time across groups for PACC (*F*(2,1015)=120.6, p<.0001), ADNI-MEM (*F*(2,1104)=55.4, p<.0001), and ADNI-EF (*F*(2,1081)=51.23, p<.0001). Estimated rates of change (slopes) for each cognitive measure are shown in **Table 3 and Figure 2b**. Across all three outcome measures, the AD group experienced the steepest decline, and the MCI group also declined but less steeply. The CU group did not experience a decline in either PACC (p=0.70) or ADNI-EF (p=0.22), but experienced a statistically significant, albeit small in magnitude, increase in ADNI-MEM (p=0.01).

**Table 3.** Estimated slopes (change in outcome for a 1 month increase in time) for cognitive outcomes from linear mixed effects models

| Outcome | Diagnostic Group | Slope (SE) | p-value |
| --- | --- | --- | --- |
| PACC | CU | 0.0034 (0.0090) | 0.70 |
|  | MCI | -0.074 (0.0071) | <.0001 |
|  | AD | -0.26 (0.014) | <.0001 |
| ADNI-MEM | CU | 0.0025 (0.0010) | 0.01 |
|  | MCI | -0.0050 (0.00081) | <.0001 |
|  | AD | -0.017 (0.0016) | <.0001 |
| ADNI-EF | CU | 0.0018 (0.0015) | 0.22 |
|  | MCI | -0.0038 (0.0012) | 0.001 |
|  | AD | -0.026 (0.0024) | <.0001 |

Adjusted for age, years of education, sex, *APOE* status (ε4 allele present vs absent)

**Figure 2:**
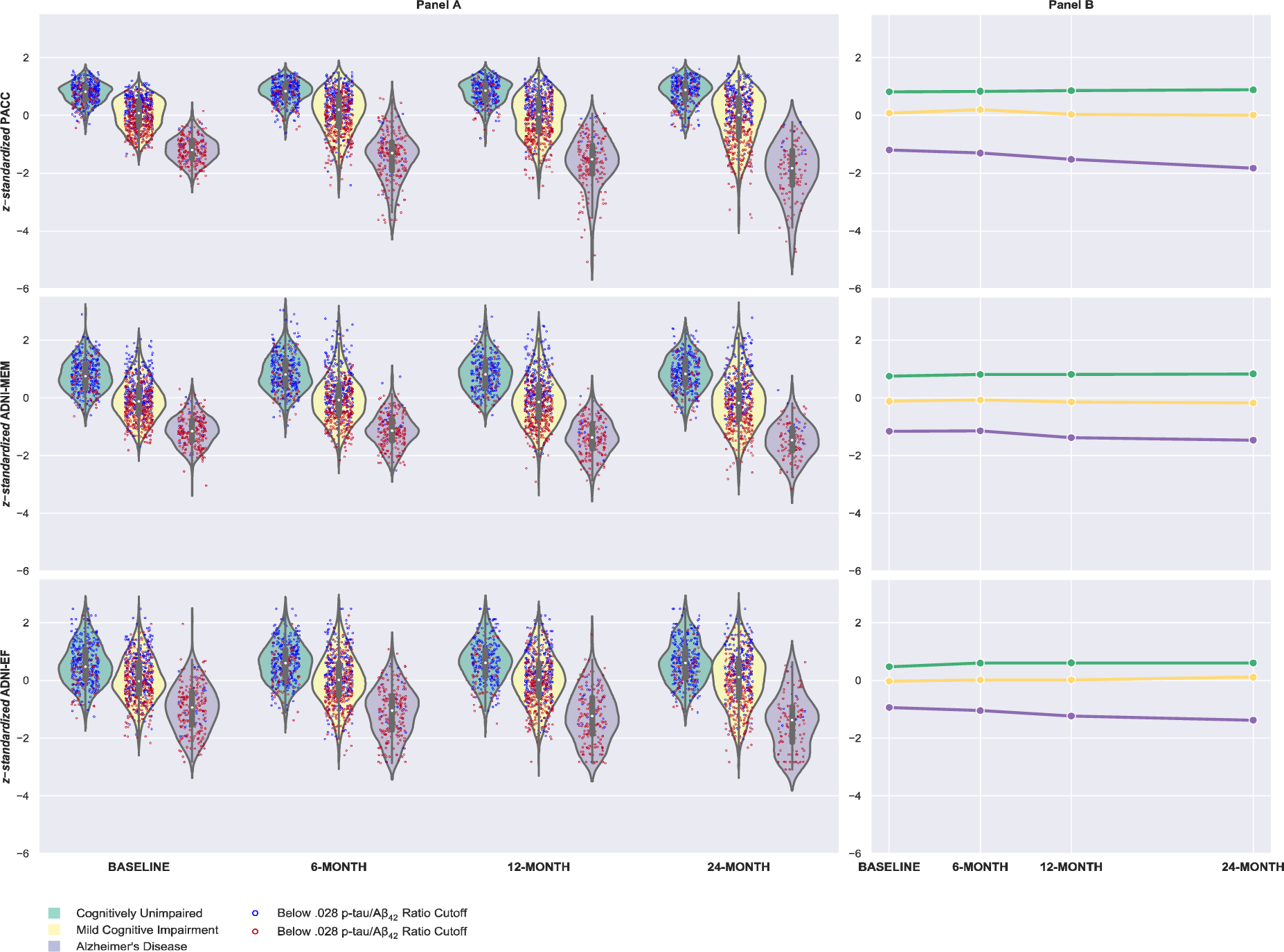
Panel A presents the distribution of z-standardized global cognition (PACC scores), episodic memory (ADNI-MEM scores), and executive functioning (ADNI-EF scores) across the three diagnostic groups, and Panel B shows the decline in the three cognitive domains across the two-year period.

### Effects of AD Pathology Biomarkers and Diagnostic Group on Metrics of Cognitive Functioning

When the p-tau/Aβ_42_ ratio was added into the mixed effects models, the three-way interaction of diagnosis group by p-tau/Aβ_42_ by time was significant for PACC (*p* = .02) and ADNI-MEM (*p* = .004) (**Table 4; Figure 3**). Contrasts in the models revealed that higher p-tau/Aβ_42_ was associated with steeper declines in PACC over time for the AD group (*p* = .001) and the MCI group (*p* < .0001), but for the CU group there was no effect of p-tau/Aβ_42_ on change over time (*p* = .51). Similarly, higher p-tau/Aβ_42_ was associated with steeper declines in ADNI-MEM for the AD group (*p* = .001) and the MCI group (*p* < .0001) but not the CU group (*p* = .27).The three-way interaction of p-tau/Aβ_42_ by diagnosis group by time was not significant for ADNI-EF (*p* = .31), but the two-way interaction p-tau/Aβ_42_ by time was significant (*p* < .0001) indicating that there was an effect of pathology on the rate of change in ADNI-EF over time, but that this effect did not significantly differ by diagnostic group. Pooled across groups, higher ratio of p-tau/Aβ_42_ was associated with a steeper decline in ADNI-EF.

**Table 4.**
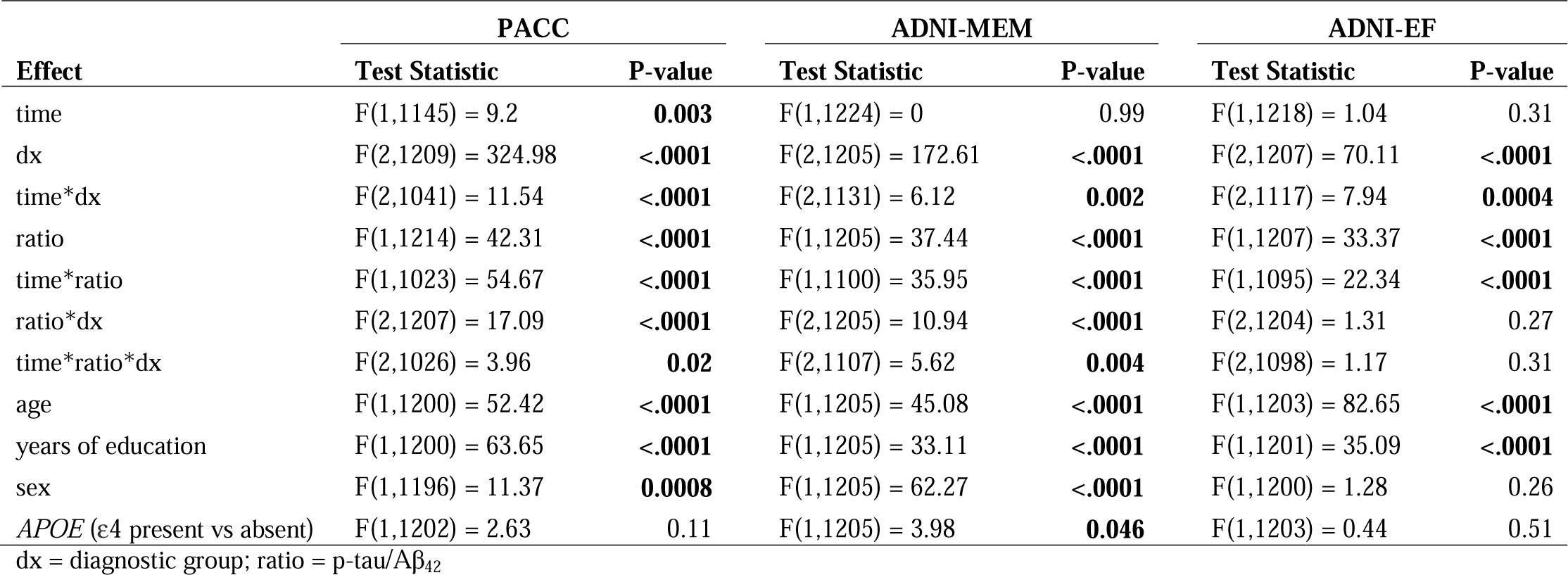
Test statistics for the linear mixed models evaluating the effect of p-tau/Aβ_42_ on cognitive outcomes.

**Figure 3:**
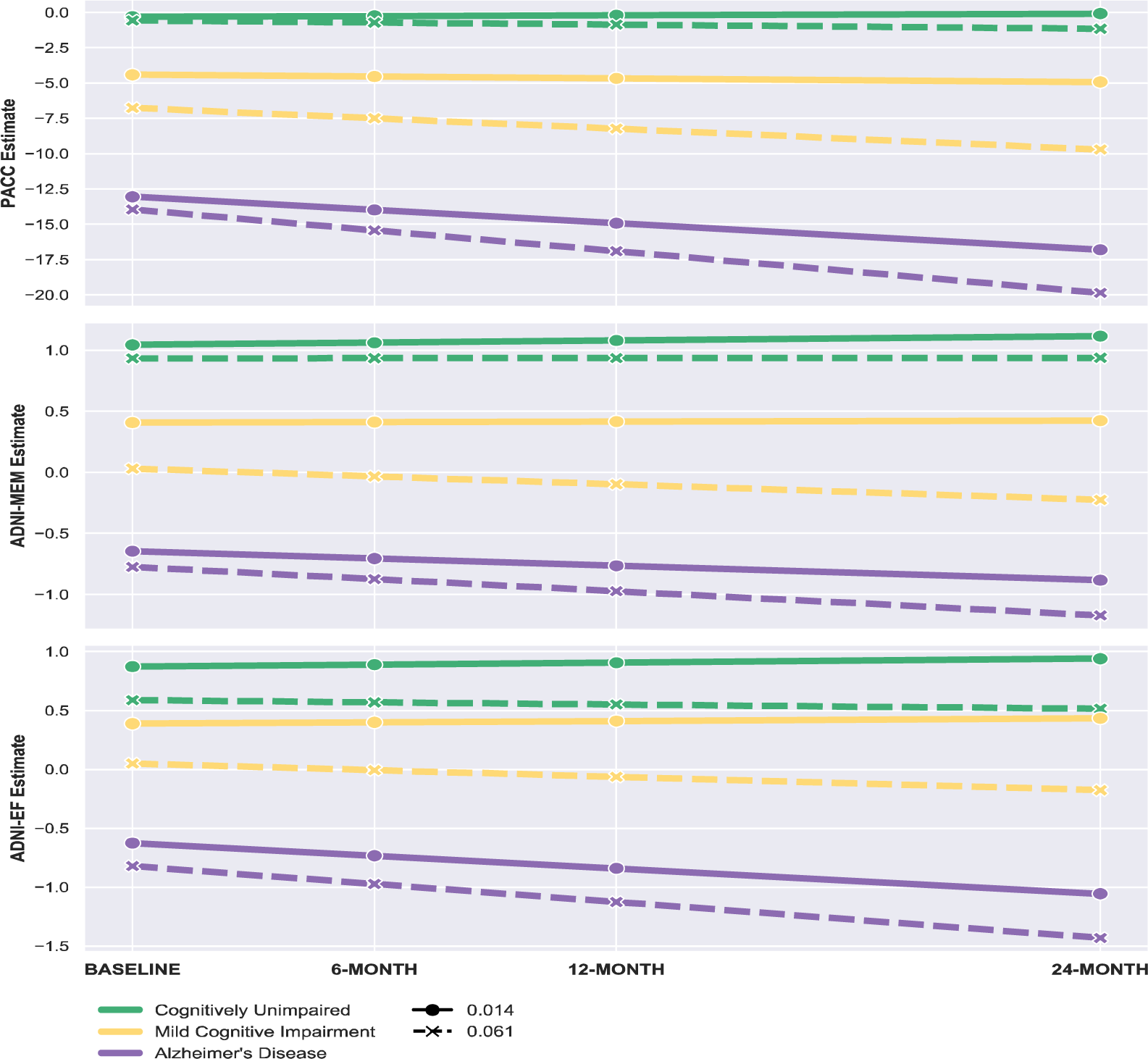
Predicted trajectories of each outcome by diagnosis group at two levels of p-tau/Aβ_42_ ratio (solid lines: ratio = 0.014 = mean of values below established cutoff; dotted lines: ratio = 0.061 = mean of values above established cutoff)

## Discussion

The conceptualization of AD on the basis of AD neuropathology includes the presence of both β-amyloid pathology and phosphyrated tau as necessary evidence for AD^11,38^. In support of this, there is growing evidence for a strong concordance between combined Aβ and tau CSF-based measures and uptake of amyloid tracers in PET imaging^26,27,29^. For example, Roe et al. examining the relationships between CSF-based and PET-based markers of pathology found that PiB uptake values were predicted by an interaction between CSF-Aβ_42_ and CSF tau^29^. In our study, as would be expected, the ratio of p-tau/Aβ_42_ was the highest in AD individuals, followed by MCI, and then CU older adults. Interestingly, the effect sizes for the CSF ratio was much larger compared to the effect size for either Aβ_42_ or p-tau, suggesting that this combined metric of CSF-based biomarkers may be best able to distinguish the three diagnostic groups employed in the study.

Besides having AD pathology, the clinical diagnosis of Alzheimer-related MCI and dementia requires clinical symptomatology, with an emphasis on multi-domain cognitive deficits including memory impairment. Importantly, AD-specific pathology, quantified here as the ratio of CSF p-tau/Aβ_42_, predicted decline in global cognition (PACC) and episodic memory (ADNI-MEM) only for individuals with MCI and AD. Decrements in memory abilities are considered to be one of the earliest signs of cognitive dysfunction related to AD^39–41^, with declines in global functioning following the memory decline, and in the later stages of the disease serving as the leading predictor of long-term cognitive changes^39,42,43^. Executive ability impairments, a broad umbrella term encompassing a multitude of top-down, prefrontal-reliant operations^44^, and known to be sensitive to age-related decline^45,46^, are also seen early on in those with AD. This study showed that a composite measure of executive functioning (ADNI-EF) was associated with CSF p-tau/Aβ_42_ across all diagnostic groups. Although the decline in cognitive functioning, and notably decrements in sub-domains of executive functioning, have been well documented with advancing age, there is significant individual variability in the temporal trajectory and scope of such deficits^46^. This study, by examining linkages between biomarkers of brain pathology and cognitive decline, provides support that the accumulation of amyloid and tau pathology worsens executive abilities not only in MCI and AD groups but also in CU older adults. This suggests that executive skills may be as early or perhaps an even earlier indicator of AD cognitive decline as memory impairments.

Our results are in agreement with the growing number of studies investigating the synergistic contribution of the two hallmark proteinopathies in predicting cognitive decline in preclinical AD. Individuals with lower levels of CSF Aβ_42_ and elevated levels of CSF p-tau have an increased risk of conversion to MCI^47^. Additionally, combined measures of amyloid beta and tau correlates with both cognitive and functional abilities. Increasing AD pathology biomarkers predicts progression on the Clinical Dementia Rating Scale^48^, cross-sectionally associates with driving performance^49^, and longitudinally predicts decline in global cognition,^32^ memory, executive functioning, and semantic fluency^33^.

Strengths of this study includes the use of composite measures to quantify cognitive domains. By employing validated composite measures that included metrics across multiple measures and reducing variability and measurement error associated with individual tasks, our measures are likely to be more sensitive at detecting hypothesized effects. Another strength is the use of continuous measures of brain pathology as predictors of cognitive decline. Prior evidence suggests sizeable heterogeneity in classification of participants based on the differences in employed biomarkers. For example, in a recent investigation from the BioFINDER study, cut-off thresholds based on CSF p-tau resulted in a larger proportion of CU participants being classified as tau-positive compared with those classified using tau PET^50^. Additionally, in the same study, sensitivities of the various biomarkers in predicting cognitive decline was substantially reduced when dichotomized values were employed compared with the use of continuous measures, especially for CU individuals where pathology accumulation is still growing.

There are a number of limitations of this study that are important to note. We included a relatively short follow-up period of two years despite availability of additional longitudinal data in the ADNI dataset. However, this a priori decision was driven based on the significant attrition in this dataset by diagnostic groups, such that there were relatively fewer number of AD participants with available follow-up data. Additionally, we limited our analyses to CSF-based biomarkers of Aβ_42_ and p-tau. It is possible that PET-based biomarkers of beta amyloid and tau provide a different pattern of results, and inclusion of other synaptic and neuronal biomarkers of AD pathophysiology could provide additional valuable insights for prediction of the clinical progression of the disease.

In conclusion, the ratio of CSF-based biomarkers of amyloid and tau pathology, show larger between-group effect sizes than the individual biomarkers when comparing CU participants with MCI and AD individuals. Worsening neuropathological changes seen in participants predicted declines in global metrics of cognition and episodic memory in those with MCI and AD and predicted declines in executive abilities in those with MCI, AD, and cognitively unimpaired participants suggesting executive impairments may occur very early in AD individuals. Amyloid and tau pathology are potential sources of heterogeneity explaining some of the variability in AD-related decline in global cognition, episodic memory, and executive functioning.

## Supporting information

ADNI Co-investigator appendix

## Data Availability

We analyzed data from the publicly available Alzheimer's Disease Neuroimaging Initiative (http://adni.loni.usc.edu).

http://adni.loni.usc.edu/data-samples/access-data/

## APPENDIX 1: Authors

| Name | Location | Contribution |
| --- | --- | --- |
| Ruchika Prakash | The Ohio State University, OH | Designed and conceptualized the study; wrote the first draft of the manuscript; and incorporated feedback from all co-authors |
| Michael McKenna | The Ohio State University, OH | Downloaded and analyzed the data; revised manuscript for intellectual content |
| Oyetunde Gbadeyan | The Ohio State University, OH | Created figures; revised manuscript for intellectual content |
| Rebecca Andridge | The Ohio State University, OH | Analyzed the data; revised manuscript for intellectual content |
| Douglas Scharre | The Ohio State University, OH | Interpreted data; revised manuscript for intellectual content |

