## Supplementary material for "p-tau/Aβ42 Ratio Associates with Cognitive Decline in Alzheimer’s disease, Mild Cognitive Impairment, and Cognitively Unimpaired Older Adults": ADNI Co-investigator appendix

Michael Weiner, MD (UC San Francisco, Principal Investigator, Executive Committee); Paul Aisen, MD (UC San Diego, ADCS PI and Director of Coordinating Center Clinical Core, Executive Committee, Clinical Core Leaders); Ronald Petersen, MD, PhD (Mayo Clinic, Rochester, Executive Committee, Clinical Core Leader); Clifford R. Jack, Jr., MD (Mayo Clinic, Rochester, Executive Committee, MRI Core Leader); William Jagust, MD (UC Berkeley, Executive Committee; PET Core Leader); John Q. Trojanowki, MD, PhD (U Pennsylvania, Executive Committee, Biomarkers Core Leader); Arthur W. Toga, PhD (USC, Executive Committee, Informatics Core Leader); Laurel Beckett, PhD (UC Davis, Executive Committee, Biostatistics Core Leader); Robert C. Green, MD, MPH (Brigham and Women’s Hospital, Harvard Medical School, Executive Committee and Chair of Data and Publication Committee); Andrew J. Saykin, PsyD (Indiana University, Executive Committee, Genetics Core Leader); John Morris, MD (Washington University St. Louis, Executive Committee, Neuropathology Core Leader); Leslie M. Shaw (University of Pennsylvania, Executive Committee, Biomarkers Core Leader); Enchi Liu, PhD (Janssen Alzheimer Immunotherapy, ADNI 2 Private Partner Scientific Board Chair); Tom Montine, MD, PhD (University of Washington) ; Ronald G. Thomas, PhD (UC San Diego); Michael Donohue, PhD (UC San Diego); Sarah Walter, MSc (UC San Diego); Devon Gessert (UC San Diego); Tamie Sather, MS (UC San Diego,); Gus Jiminez, MBS (UC San Diego); Danielle Harvey, PhD (UC Davis;); Michael Donohue, PhD (UC San Diego); Matthew Bernstein, PhD (Mayo Clinic, Rochester); Nick Fox, MD (University of London); Paul Thompson, PhD (USC School of Medicine); Norbert Schuff, PhD (UCSF MRI); Charles DeCArli, MD (UC Davis); Bret Borowski, RT (Mayo Clinic); Jeff Gunter, PhD (Mayo Clinic); Matt Senjem, MS (Mayo Clinic); Prashanthi Vemuri, PhD (Mayo Clinic); David Jones, MD (Mayo Clinic); Kejal Kantarci (Mayo Clinic); Chad Ward (Mayo Clinic); Robert A. Koeppe, PhD (University of Michigan, PET Core Leader); Norm Foster, MD (University of Utah); Eric M. Reiman, MD (Banner Alzheimer’s Institute); Kewei Chen, PhD (Banner Alzheimer’s Institute); Chet Mathis, MD (University of Pittsburgh); Susan Landau, PhD (UC Berkeley); Nigel J. Cairns, PhD, MRCPath (Washington University St. Louis); Erin Householder (Washington University St. Louis); Lisa Taylor Reinwald, BA, HTL (Washington University St. Louis); Virginia Lee, PhD, MBA (UPenn School of Medicine); Magdalena Korecka, PhD (UPenn School of Medicine); Michal Figurski, PhD (UPenn School of Medicine); Karen Crawford (USC); Scott Neu, PhD (USC); Tatiana M. Foroud, PhD (Indiana University); Steven Potkin, MD UC (UC Irvine); Li Shen, PhD (Indiana University); Faber Kelley, MS, CCRC (Indiana University); Sungeun Kim, PhD (Indiana University); Kwangsik Nho, PhD (Indiana University); Zaven Kachaturian, PhD (Khachaturian, Radebaugh & Associates, Inc and Alzheimer’s Association’s Ronald and Nancy Reagan’s Research Institute); Richard Frank, MD, PhD (General Electric); Peter J. Snyder, PhD (Brown University); Susan Molchan, PhD (National Institute on Aging/ National Institutes of Health); Jeffrey Kaye, MD (Oregon Health and Science University); Joseph Quinn, MD (Oregon Health and Science University); Betty Lind, BS (Oregon Health and Science University); Raina Carter, BA (Oregon Health and Science University); Sara Dolen, BS (Oregon Health and Science University); Lon S. Schneider, MD (University of Southern California); Sonia Pawluczyk, MD (University of Southern California); Mauricio Beccera, BS (University of Southern California); Liberty Teodoro, RN (University of Southern California); Bryan M. Spann, DO, PhD (University of Southern California); James Brewer, MD, PhD (University of California San Diego); Helen Vanderswag, RN (University of California San Diego); Adam Fleisher, MD (University of California San Diego); Judith L. Heidebrink, MD, MS (University of Michigan); Joanne L. Lord, LPN, BA, CCRC (University of Michigan); Ronald Petersen, MD, PhD (Mayo Clinic, Rochester); Sara S. Mason, RN (Mayo Clinic, Rochester); Colleen S. Albers, RN (Mayo Clinic, Rochester); David Knopman, MD (Mayo Clinic, Rochester); Kris Johnson, RN (Mayo Clinic, Rochester); Rachelle S. Doody, MD, PhD (Baylor College of Medicine); Javier Villanueva Meyer, MD (Baylor College of Medicine); Munir Chowdhury, MBBS, MS (Baylor College of Medicine); Susan Rountree, MD (Baylor College of Medicine); Mimi Dang, MD (Baylor College of Medicine); Yaakov Stern, PhD (Columbia University Medical Center); Lawrence S. Honig, MD, PhD (Columbia University Medical Center); Karen L. Bell, MD (Columbia University Medical Center); Beau Ances, MD (Washington University, St. Louis); John C. Morris, MD (Washington University, St. Louis); Maria Carroll, RN, MSN (Washington University, St. Louis); Sue Leon, RN, MSN (Washington University, St. Louis); Erin Householder, MS, CCRP (Washington University, St. Louis); Mark A. Mintun, MD (Washington University, St. Louis); Stacy Schneider, APRN, BC, GNP (Washington University, St. Louis); Angela Oliver, RN, BSN, MSG ; Daniel Marson, JD, PhD (University of Alabama Birmingham); Randall Griffith, PhD, ABPP (University of Alabama Birmingham); David Clark, MD (University of Alabama Birmingham); David Geldmacher, MD (University of Alabama Birmingham); John Brockington, MD (University of Alabama Birmingham); Erik Roberson, MD (University of Alabama Birmingham); Hillel Grossman, MD (Mount Sinai School of Medicine); Effie Mitsis, PhD (Mount Sinai School of Medicine); Leyla deToledo-Morrell, PhD (Rush University Medical Center); Raj C. Shah, MD (Rush University Medical Center); Ranjan Duara, MD (Wien Center); Daniel Varon, MD (Wien Center); Maria T. Greig, HP (Wien Center); Peggy Roberts, CNA (Wien Center); Marilyn Albert, PhD (Johns Hopkins University); Chiadi Onyike, MD (Johns Hopkins University); Daniel D’Agostino II, BS (Johns Hopkins University); Stephanie Kielb, BS (Johns Hopkins University); James E. Galvin, MD, MPH (New York University); Dana M. Pogorelec (New York University); Brittany Cerbone (New York University); Christina A. Michel (New York University); Henry Rusinek, PhD (New York University); Mony J de Leon, EdD (New York University); Lidia Glodzik, MD, PhD (New York University); Susan De Santi, PhD (New York University); P. Murali Doraiswamy, MD (Duke University Medical Center); Jeffrey R. Petrella, MD (Duke University Medical Center); Terence Z. Wong, MD (Duke University Medical Center); Steven E. Arnold, MD (University of Pennsylvania); Jason H. Karlawish, MD (University of Pennsylvania); David Wolk, MD (University of Pennsylvania); Charles D. Smith, MD (University of Kentucky); Greg Jicha, MD (University of Kentucky); Peter Hardy, PhD (University of Kentucky); Partha Sinha, PhD (University of Kentucky); Elizabeth Oates, MD (University of Kentucky); Gary Conrad, MD (University of Kentucky); Oscar L. Lopez, MD (University of Pittsburgh); MaryAnn Oakley, MA (University of Pittsburgh); Donna M. Simpson, CRNP, MPH (University of Pittsburgh); Anton P. Porsteinsson, MD (University of Rochester Medical Center); Bonnie S. Goldstein, MS, NP (University of Rochester Medical Center); Kim Martin, RN (University of Rochester Medical Center); Kelly M. Makino, BS (University of Rochester Medical Center); M. Saleem Ismail, MD (University of Rochester Medical Center); Connie Brand, RN (University of Rochester Medical Center); Ruth A. Mulnard, DNSc, RN, FAAN (University of California, Irvine); Gaby Thai, MD (University of California, Irvine); Catherine Mc Adams Ortiz, MSN, RN, A/GNP (University of California, Irvine); Kyle Womack, MD (University of Texas Southwestern Medical School); Dana Mathews, MD, PhD (University of Texas Southwestern Medical School); Mary Quiceno, MD (University of Texas Southwestern Medical School); Ramon Diaz Arrastia, MD, PhD (University of Texas Southwestern Medical School); Richard King, MD (University of Texas Southwestern Medical School); Myron Weiner, MD (University of Texas Southwestern Medical School); Kristen Martin Cook, MA (University of Texas Southwestern Medical School); Michael DeVous, PhD (University of Texas Southwestern Medical School); Allan I. Levey, MD, PhD (Emory University); James J. Lah, MD, PhD (Emory University); Janet S. Cellar, DNP, PMHCNS BC (Emory University); Jeffrey M. Burns, MD (University of Kansas, Medical Center); Heather S. Anderson, MD (University of Kansas, Medical Center); Russell H. Swerdlow, MD (University of Kansas, Medical Center); Liana Apostolova, MD (University of California, Los Angeles); Kathleen Tingus, PhD (University of California, Los Angeles); Ellen Woo, PhD (University of California, Los Angeles); Daniel H.S. Silverman, MD, PhD (University of California, Los Angeles); Po H. Lu, PsyD (University of California, Los Angeles); George Bartzokis, MD (University of California, Los Angeles); Neill R Graff Radford, MBBCH, FRCP (London) (Mayo Clinic, Jacksonville); Francine Parfitt, MSH, CCRC (Mayo Clinic, Jacksonville); Tracy Kendall, BA, CCRP (Mayo Clinic, Jacksonville); Heather Johnson, MLS, CCRP (Mayo Clinic, Jacksonville); Martin R. Farlow, MD (Indiana University); Ann Marie Hake, MD (Indiana University); Brandy R. Matthews, MD (Indiana University); Scott Herring, RN, CCRC (Indiana University); Cynthia Hunt, BS, CCRP (Indiana University); Christopher H. van Dyck, MD (Yale University School of Medicine); Richard E. Carson, PhD (Yale University School of Medicine); Martha G. MacAvoy, PhD (Yale University School of Medicine); Howard Chertkow, MD (McGill Univ., Montreal Jewish General Hospital); Howard Bergman, MD (McGill Univ., Montreal Jewish General Hospital); Chris Hosein, Med (McGill Univ., Montreal Jewish General Hospital); Sandra Black, MD, FRCPC (Sunnybrook Health Sciences, Ontario); Dr Bojana Stefanovic (Sunnybrook Health Sciences, Ontario); Curtis Caldwell, PhD (Sunnybrook Health Sciences, Ontario); Ging Yuek Robin Hsiung, MD, MHSc, FRCPC (U.B.C. Clinic for AD & Related Disorders); Howard Feldman, MD, FRCPC (U.B.C. Clinic for AD & Related Disorders); Benita Mudge, BS (U.B.C. Clinic for AD & Related Disorders); Michele Assaly, MA Past (U.B.C. Clinic for AD & Related Disorders); Andrew Kertesz, MD (Cognitive Neurology St. Joseph’s, Ontario); John Rogers, MD (Cognitive Neurology St. Joseph’s, Ontario); Dick Trost, PhD (Cognitive Neurology St. Joseph’s, Ontario); Charles Bernick, MD (Cleveland Clinic Lou Ruvo Center for Brain Health); Donna Munic, PhD (Cleveland Clinic Lou Ruvo Center for Brain Health); Diana Kerwin, MD (Northwestern University); Marek Marsel Mesulam, MD (Northwestern University); Kristine Lipowski, BA (Northwestern University); Chuang Kuo Wu, MD, PhD (Northwestern University); Nancy Johnson, PhD (Northwestern University); Carl Sadowsky, MD (Premiere Research Inst (Palm Beach Neurology)); Walter Martinez, MD (Premiere Research Inst (Palm Beach Neurology)); Teresa Villena, MD (Premiere Research Inst (Palm Beach Neurology)); Raymond Scott Turner, MD, PhD (Georgetown University Medical Center); Kathleen Johnson, NP (Georgetown University Medical Center); Brigid Reynolds, NP (Georgetown University Medical Center); Reisa A. Sperling, MD (Brigham and Women’s Hospital); Keith A. Johnson, MD (Brigham and Women’s Hospital); Gad Marshall, MD (Brigham and Women’s Hospital); Meghan Frey (Brigham and Women’s Hospital); Jerome Yesavage, MD (Stanford University); Joy L. Taylor, PhD (Stanford University); Barton Lane, MD (Stanford University); Allyson Rosen, PhD (Stanford University); Jared Tinklenberg, MD (Stanford University); Marwan N. Sabbagh, MD (Banner Sun Health Research Institute); Christine M. Belden, PsyD (Banner Sun Health Research Institute); Sandra A. Jacobson, MD (Banner Sun Health Research Institute); Sherye A. Sirrel, MS (Banner Sun Health Research Institute); Neil Kowall, MD (Boston University); Ronald Killiany, PhD (Boston University); Andrew E. Budson, MD (Boston University); Alexander Norbash, MD (Boston University); Patricia Lynn Johnson, BA (Boston University); Thomas O. Obisesan, MD, MPH (Howard University); Saba Wolday, MSc (Howard University); Joanne Allard, PhD (Howard University); Alan Lerner, MD (Case Western Reserve University); Paula Ogrocki, PhD (Case Western Reserve University); Leon Hudson, MPH (Case Western Reserve University); Evan Fletcher, PhD (University of California, Davis Sacramento); Owen Carmichael, PhD (University of California, Davis Sacramento); John Olichney, MD (University of California, Davis Sacramento); Charles DeCarli, MD (University of California, Davis Sacramento); Smita Kittur, MD (Neurological Care of CNY); Michael Borrie, MB ChB (Parkwood Hospital); T Y Lee, PhD (Parkwood Hospital); Dr Rob Bartha, PhD (Parkwood Hospital); Sterling Johnson, PhD (University of Wisconsin); Sanjay Asthana, MD (University of Wisconsin); Cynthia M. Carlsson, MD (University of Wisconsin); Steven G. Potkin, MD (University of California, Irvine BIC); Adrian Preda, MD (University of California, Irvine BIC); Dana Nguyen, PhD (University of California, Irvine BIC); Pierre Tariot, MD (Banner Alzheimer’s Institute); Adam Fleisher, MD (Banner Alzheimer’s Institute); Stephanie Reeder, BA (Banner Alzheimer’s Institute); Vernice Bates, MD (Dent Neurologic Institute); Horacio Capote, MD (Dent Neurologic Institute); Michelle Rainka, PharmD, CCRP (Dent Neurologic Institute); Douglas W. Scharre, MD (Ohio State University); Maria Kataki, MD, PhD (Ohio State University); Anahita Adeli, MD (Ohio State University); Earl A. Zimmerman, MD (Albany Medical College); Dzintra Celmins, MD (Albany Medical College); Alice D. Brown, FNP (Albany Medical College); Godfrey D. Pearlson, MD (Hartford Hosp, Olin Neuropsychiatry Research Center); Karen Blank, MD (Hartford Hosp, Olin Neuropsychiatry Research Center); Karen Anderson, RN (Hartford Hosp, Olin Neuropsychiatry Research Center); Robert B. Santulli, MD (Dartmouth Hitchcock Medical Center); Tamar J. Kitzmiller (Dartmouth Hitchcock Medical Center); Eben S. Schwartz, PhD (Dartmouth Hitchcock Medical Center); Kaycee M. Sink, MD, MAS (Wake Forest University Health Sciences); Jeff D. Williamson, MD, MHS (Wake Forest University Health Sciences); Pradeep Garg, PhD (Wake Forest University Health Sciences); Franklin Watkins, MD (Wake Forest University Health Sciences); Brian R. Ott, MD (Rhode Island Hospital); Henry Querfurth, MD (Rhode Island Hospital); Geoffrey Tremont, PhD (Rhode Island Hospital); Stephen Salloway, MD, MS (Butler Hospital); Paul Malloy, PhD (Butler Hospital); Stephen Correia, PhD (Butler Hospital); Howard J. Rosen, MD (UC San Francisco); Bruce L. Miller, MD (UC San Francisco); Jacobo Mintzer, MD, MBA (Medical University South Carolina); Kenneth Spicer, MD, PhD (Medical University South Carolina); David Bachman, MD (Medical University South Carolina); Elizabether Finger, MD (St. Joseph’s Health Care); Stephen Pasternak, MD (St. Joseph’s Health Care); Irina Rachinsky, MD (St. Joseph’s Health Care); John Rogers, MD (St. Joseph’s Health Care); Andrew Kertesz, MD (St. Joseph’s Health Care); Dick Drost, MD (St. Joseph’s Health Care); Nunzio Pomara, MD (Nathan Kline Institute); Raymundo Hernando, MD (Nathan Kline Institute); Antero Sarrael, MD (Nathan Kline Institute); Susan K. Schultz, MD (University of Iowa College of Medicine, Iowa City); Laura L. Boles Ponto, PhD (University of Iowa College of Medicine, Iowa City); Hyungsub Shim, MD (University of Iowa College of Medicine, Iowa City); Karen Elizabeth Smith, RN (University of Iowa College of Medicine, Iowa City); Norman Relkin, MD, PhD (Cornell University); Gloria Chaing, MD (Cornell University); Lisa Raudin, PhD (Cornell University); Amanda Smith, MD (University of South Florida: USF Health Byrd Alzheimer’s Institute); Kristin Fargher, MD (University of South Florida: USF Health Byrd Alzheimer’s Institute); Balebail Ashok Raj, MD (University of South Florida: USF Health Byrd Alzheimer’s Institute)
